# Objective and Subjective Assessments of Exercise Burden in Masters Athletes Are Poorly Correlated

**DOI:** 10.1101/2023.12.06.23299458

**Authors:** Jennifer Lewis, Robert F. Bentley, Kim A. Connelly, Paul Dorian, Jack M. Goodman

**Author notes:** Mailing Address (for Jack Goodman): 55 Harbord Street Toronto, Ontario M5S 2W6 Canada Email Address (for Jack Goodman). **Institution where work was performed:** University of Toronto, Toronto, Ontario, Canada.

## Abstract

Accurate quantification of exercise volume (burden) is crucial for understanding links between exercise and cardiovascular outcomes in older endurance athletes (EA). Exercise burden, an integral of intensity and duration (MET·min), is typically determined from subjective self-reports but has uncertain accuracy. We studied 40 EAs (41 to 69 yrs., 50% female) with >10 yrs. training history, during a typical outdoor cycling training session (42 km). Subjective self-reports were related to cardiac (HR·min) and metabolic (MET·min) components of exercise burden, monitored continuously. Subjective self-reports were highly variable and underestimated objective metrics of exercise intensity. Discordance was observed between metabolic and cardiac burden as less fit individuals accrued greater cardiac (14039±2649 vs. 11784±1132 HR·min*, P*<0.01) but lower metabolic burden (808±59 vs. 858±61 MET·min, *P*<0.05) vs. higher fit EA. Caution is advised in interpreting MET·min estimates from self-reports, urging objective measurement of cardiac burden for further insights into the risk-benefit relationship of long-term exercise.

## INTRODUCTION

Sustained adherence to physical activity recommendations (1, 2) is associated with a lower risk of all-cause mortality and various chronic diseases (3–5), including cardiovascular illness (6). There is increasing benefit as exercise volume increases, yet there is some evidence (7, 8), which is in dispute (6), that suggests prolonged high-intensity exercise may increase the risk of adverse cardiovascular outcomes. The relationship between exercise volume and in particular, exercise intensity, and cardiovascular disease or other health outcomes is not completely understood.

Studies examining the cumulative effects of exercise dose on cardiovascular outcomes have been largely based on non-athletic populations, relying on self-reported accounts of exercise frequency, duration, and intensity, all of which contribute to the overall exercise ‘burden’, despite uncertain accuracy. However, subjective estimates of exercise intensity have been poorly correlated to objective measures of intensity due to recall bias and confounding factors including fitness level, BMI, and sex (9, 10). When considering the exercise dose-cardiovascular response relationship in endurance athletes, studies have often failed to directly assess exercise intensity and more importantly, distinguish between the overall *metabolic* (i.e., oxygen consumption) and *cardiac-*specific (i.e., heart rate) components of the exercise dose-response relationship. This complex relationship is predicated on accurate quantification of exercise intensity, a key determining factor determining cardiovascular adaptations to exercise training (11) and the cumulative exercise ‘volume’ associated with an elevated risk for certain adverse outcomes (12). These considerations are particularly pertinent for older endurance athletes with long-standing exercise history who reflect the largest and fastest growing cohort of mass participation events (13), and who may have increased cardiovascular disease burden, especially an elevated risk for developing lone atrial fibrillation (14). Without an accurate determination of exercise intensity, reports of exercise histories may be inaccurate and misleading, particularly when attributing high levels of cumulative exercise ‘burden’ to adverse long-term cardiovascular outcomes.

The purpose of this study was to assess subjective reports of exercise intensity during typical endurance training in the field and relate these to objective measures of intensity. We sought to determine if estimates of exercise intensity from self-reports would accurately reflect objective measures of intensity in masters endurance athletes.

## METHODS

### Participants

Male and female adults aged 40 to 69 years were recruited from local cycling clubs. Inclusion criteria included experience with standard road cycling including a weekly ride of ≥60 km and total weekly mileage of ≥100 km. Exclusion criteria included ranking as current or former national or Olympic team cyclists, history of smoking, cardiovascular disease, metabolic disorders, hypertension with resting pressures exceeding 140/90 despite medication (mmHg), sleep apnea, recent infection or inflammation, and thyroid disease. Participants completed a written informed consent approved by the University of Toronto Health Sciences Research Ethics Board (Protocol #39300) which included directives and restrictions related to the COVID-19 pandemic at the time (October-November 2020). The study was conducted in full compliance with the Declaration of Helsinki and its amendments.

### Field Study Exercise Protocol

This prospective observational cohort field study was designed to mimic a typical mid-distance, self-paced training session on a 42 km pre-determined route in a rural setting. Participants were instructed to ride at a ‘typical’ training intensity along a pre-determined cycling route on side roads of varying topography that would elicit different levels of challenge (345 m elevation change); pre-determined landmarks identified where ratings of perceived exertion (RPE) were obtained via wireless communication during brief sections (2 to 3 km distance) of ’self-paced’ efforts at prespecified ‘low’, ‘medium’, ‘high’ and ‘very high’ levels of effort (Figure 1). All participants abstained from alcohol, caffeine, and endurance training for at least 24 hours prior to the field study.

**Figure 1.**
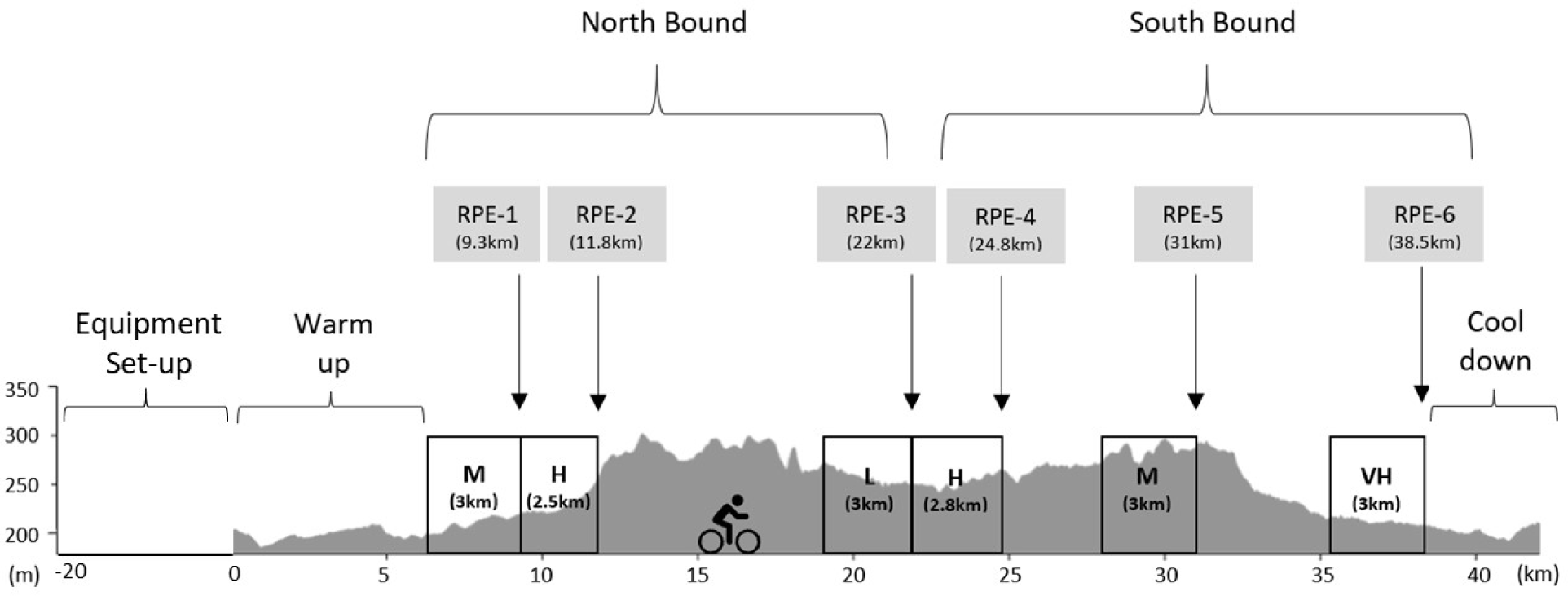
Course profile and rating-of-perceived-exertion (RPE) landmarks. Prescribed efforts: L=low effort, M=medium effort, H=high effort, VH=very high effort.

### Baseline Measures and Preparation for Exercise

Upon arrival at the staging area before the ride, participants were provided with a detailed familiarization session and completed a questionnaire (15) assessing the quality of their previous night’s sleep, current state of fatigue, stress, and muscle soreness. Body mass (kg) was assessed using a digital scale (Starfrit Balance, Atlantic Promotions Inc., Canada), and resting blood pressure (BP) and resting heart rate (HR) were obtained from three consecutive measures from a BP monitor (Omron 10 Series, Model BP7450CAN, Canada). Height (cm) was self-reported.

Participants were then refamiliarized with RPE scales, including the Borg 6-20 (RPE_Borg_) (16) and the Word scale (RPE_Word_) (17), where: 1=very light, 2=light, 3=moderate, 4=vigorous, 5=extremely vigorous. Participants were then fitted with equipment that provided continual measurements of heart rate and work rate which began at rest for 5 minutes in a seated position. They were also fitted with a portable earphone. Bike and communication set-up were tested by the participants in the staging area before ride departure.

### Exercise Protocol and Monitoring

Participants performed the ride at their preferred cadence and training effort until called upon to complete the prespecified paced session (Figure 1) and then verbally reported their subjective effort using both RPE scales, in random order, without feedback provided. Following the completion of the exercise, a measure of the global perception of effort for the entire ride (post-ride RPE) was obtained 30 minutes after recovery using the Borg 6-20 and Word scales.

The warm-up (first 6 km) and cool-down (last 3.5 km) during the route were performed as per their usual routine, and non-caffeinated fluid and fuel ingestion were permitted, ad libitum.

Due to COVID-19-related face-to-face research restrictions, direct laboratory assessment or in-field measures of VO_2max_ were not possible, therefore VO_2peak_ was estimated from peak power and heart rate data. The maximal heart rate observed was derived from the average of two consecutive 60-sec recordings, compared against age-predicted HR_max_ (18), and VO_2peak_ was then estimated based on the peak power aligned to these values (19).

Communication (for monitoring and soliciting RPE scores) between the investigator and participants utilized hands-free, automatic call-answering communication through a mobile device (s10 or s20 Plus, Samsung Electronics Co. Ltd., South Korea) mounted on the bike with a portable earphone (AirPods Pro, Apple Inc., USA) secured before starting the ride. Remote tracking of participants was provided by asset tracking software (Fluid Mobility Inc., Ontario, Canada), and geo-fencing ensured all participants were at the same location when reporting RPE.

### Physiological Monitoring and Data Processing

Heart rate (HR) was continuously monitored using a Viiiiva chest strap (4iiii Innovations Inc., Alberta, Canada) and a Frontier X chest strap (Frontier X, Fourth Frontier Technologies Private Limited, Bangalore, India). Participants’ road bikes were fitted with calibrated power meter pedals (Garmin Vector 3, Garmin International Inc., Olathe, KS, USA) and Vector cleats (Arc R2, Garmin, USA). The Viiiiva heart rate monitor and Vector 3 pedals were synced with a portable computer (Edge 530, Garmin International Inc., Olathe, KS, USA), allowing for continuous monitoring of power (watts), with only speed, time and direction measures visible to the participants. All measures were recorded continuously throughout the ride for each device at sampling frequencies of 125 Hz (HR) or 1 Hz (power). Heart rate and power data were averaged over one-minute intervals. Age-predicted maximal heart rate was calculated using Tanaka’s formula (18), and percentage of effort for HR and METs was relative to the peak levels achieved during the ride. RPE was recorded at six distinct landmarks (LM1 to LM6). The first 120s and last 60s of each ride were excluded from the analysis to account for variations in departure and arrival routines.

The total exercise cardiac burden of the field ride was determined by an area under the curve (AUC) analysis for individual participants’ heart rate data, obtained from the continuous recordings averaged over 1-minute intervals data for (AUC HR, expressed as HR·min). This measure is an estimate of the total number of heartbeats during the ride. Similarly, the estimated total metabolic burden (MET·min) of the entire ride was derived from AUC analysis calculated from the average power (watts) and body mass (kg) over time, using the ACSM equation for oxygen cost of cycling/leg ergometry (19). Total MET·min is an estimate of (body mass corrected) total energy produced during the ride.

### Statistical Analysis

Normality of data was assessed with a Shapiro-Wilk test. All normally distributed data are reported as mean ± standard deviation while non-normally distributed data are median (interquartile range). To explore the effect of each landmark on heart rate, power, and calculated METs, one-way repeated measures ANOVAs were completed. When the assumption of sphericity was violated, a Greenhouse-Geisser correction was applied. Following a significant landmark effect, Bonferroni corrected *post hoc* tests were completed. To explore the effect of biological sex on objective and subjective measures over the entire ride and participant characteristics, independent samples t-tests and Mann-Whitney U tests were completed as appropriate. The effect of fitness on ride endpoints was assessed with independent samples t-tests and Mann-Whitney U tests as appropriate. The associations between cardiac and metabolic burden and ride parameters were assessed with Pearson correlations and Spearman Rho as appropriate. Statistical significance was set at a two-tailed alpha level of 0.05. All statistical analyses were carried out using SPSS Statistics software 26 (IBM Corporation, Armonk, NY, USA), and AUC calculations used SigmaPlot 11 graphing software (Systat Software Inc., California, USA).

## RESULTS

Forty masters athletes (50% female) between 41 and 69 years of age completed the study. Baseline characteristics are summarized in Table 1. Participants had an average history of recreational and or competitive training for cycling equal to 15±9 years or mixed endurance training equal to 19±2 years. All reported normal sleep patterns and duration (*median*=7.6 hours) the night before testing and did not report unusual muscle soreness, fatigue, or stress.

**Table 1.** Participant characteristics.

|  | Males (n=20) | Females (n=20) | Group (n=40) |
| --- | --- | --- | --- |
| Age (years) | 55 ± 8 | 53 ± 7 | 54 ± 8 |
| Body Mass (kg) | 80.3 ± 7.9** | 63.2 ± 9.5 | 71.7 ± 12.2 |
| Height (cm) | 178 ± 7** | 166 ± 8 | 172 ± 10 |
| BMI (kg/m<sup>2</sup>) | 25.3 ± 2.8** | 22.8 ± 2.9 | 24.1 ± 3.1 |
| BSA (m<sup>2</sup>) | 1.99 ± 0.11** | 1.71 ± 0.16 | 1.85 ± 0.20 |
| Resting HR (bpm) | 60 ± 13 | 63 ± 9 | 62 ± 11 |
| Resting SBP (mmHg) | 140 ± 17* | 128 ± 17 | 134 ± 18 |
| Resting DBP (mmHg) | 84 ± 11 | 81 ± 12 | 82 ± 11 |
| Estimated VO<sub>2peak</sub> (mL/kg/min) | 44.2 ± 6.6 | 40.2 ± 5.6 | 42.3 ± 6.3 |
| Prior Smoker | 20% (4) | 15% (3) | 18% (7) |
| Alcohol (drinks/week) | 5 ± 5 | 5 ± 4 | 5 ± 4 |
| Yearly Cycling Mileage (km) | 8196 ± 3837* | 5530 ± 2831 | 6863 ± 3592 |
| Weekly Cycling Duration (hours) | 9.4 ± 3.7* | 7.1 ± 3.4 | 8.2 ± 3.7 |

*Note.* Data are means ± standard deviations or percentages (number of participants). BMI, body mass index; BSA, body surface area; HR, heart rate (beats per minute); SBP, systolic blood pressure; DBP, diastolic blood pressure; estimated VO<sub>2peak</sub>, estimated peak oxygen consumption.
\* Statistically significant biological sex difference at $p < 0.05$ ; \*\* $p < 0.01$ .

### Field Conditions

Participants completed the 42 km ride without consequence during the morning (n=37) or early afternoon (n=3). Environmental conditions (Environment and Climate Change Canada) varied by ambient temperature (5 to 21°C), wind (2 to 25 km/h; gusts 0 to 44 km/h), and relative humidity (31 to 99%). Light rain occurred on one day (impacting two female riders).

### Exercise Intensity

Continuous measures of exercise intensity from the entire ride are presented in Table 2. The mean time to complete the ride was 90.0±8.6 min, at a mean cadence of 82±6 rpm, and a mean speed of 28.9±2.5 km/h, with an average power output of 173±42 W. The mean HR_peak_ achieved during the ride was 167±10 bpm, equivalent to 98% of the age-predicted HR_max_ (170±5 bpm) using Tanaka’s formula (18).

**Table 2.** Mean objective and median subjective measures for the entire ride (42 km).

|  | Males (n=20) | Females (n=20) | Group (n=40) |
| --- | --- | --- | --- |
| HR (bpm) | 143 ± 13 | 146 ± 11 | 144 ± 12 |
| WR (watts) | 202 ± 37** | 145 ± 24 | 173 ± 42 |
| WR/body mass (watts/kg) | 2.5 ± 0.4 | 2.3 ± 0.4 | 2.4 ± 0.4 |
| METs | 9.8 ± 1.2 | 9.1 ± 1.2 | 9.5 ± 1.3 |
| MET·min | 842.7 ± 64.3 | 840.9 ± 67.7 | 841.8 ± 65.2 |
| Duration (min) | 87.1 ± 7.6* | 93.0 ± 8.8 | 90.0 ± 8.6 |
| Combined Landmark RPE<sub>Borg</sub> (6-20) | 14.5 (1.1) | 14.3 (0.9) | 14.5 (1.0) |
| Combined Landmark RPE<sub>Word</sub> (1-5) | 3.6 (0.3) | 3.5 (0.2) | 3.5 (0.2) |

*Note.* Data are means ± stand deviations or medians (interquartile ranges). HR, heart rate; WR, power; METs, estimated metabolic equivalents; RPE, rate of perceived exertion; Borg, Borg Scale; Word, Word Scale.
\* Statistically significant at $p < 0.05$ , two-tailed; \*\* $p < 0.01$ , two-tailed.

The mean relative percentage of effort (relative to peak levels achieved) during the entire ride for heart rate and METs was 87±4% (range 80-98%) and 79±6% (range 64-101%), respectively. Mean power output was significantly higher in males than in females, but similar relative intensities were observed as there were no differences observed between biological sex for %HR_peak_ or %VO_2peak_.

Predicted %HR_max_ achieved and total duration for the ride were positively correlated (r_s_=0.36, 95% CI 0.23 to 0.71, *P*=0.02), whereas a negative correlation was observed between ride duration and mean METs (r_s_=-0.82, 95% CI -0.90 to -0.69, *P*<0.001) and estimated VO_2peak_ (r_s_=-0.79, 95% CI -0.88 to -0.64, *P*<0.001). Absolute and relative heart rate and power data at each landmark are presented in Supplementary Data Table 1. The correlation between mean power output (watts) and mean heart rate from the landmarks was r_s_=0.34 (*P*=0.001). Each measure was obtained within 0.35±0.06 km of each pre-determined landmark location. The mean MET level (gross, including resting energy) corresponding to each landmark ranged considerably from 7.7 to 12.1 METs (Suppl Table 1).

### Metabolic and Cardiac Burden: Area Under the Curve (AUC) Analyses

The calculated AUC power throughout the exercise was higher for males versus females (17,185±2253 versus 13,155±1580 W·min, *P*=0.001), but differences were not apparent in AUC power when controlling for body mass between groups (Males: 214±22 W·kg^-1^·min; Females: 210±24 W·kg^-1^·min, *P*=0.57). Males had a lower AUC HR [12,109 (2099) bpm·min] compared with females [13,320 (2960) bpm·min] (U=127.00, z=-1.975, *P*=0.048).

When participants were grouped by faster (median ≤ 89.0 min) versus slower (median >89.0 min) finishing times, the faster group had a lower AUC HR [11,711.5 (1556.8) bpm·min versus those with slower finishing times (AUC HR [13,856.5 (2043.4) bpm·min] (U=42.00, z=-4.272, *P*=0.001). However, there was no difference between the two groups for AUC METs [Faster group: 846 (84) MET·min; Slower group: 808 (96) MET·min] (U=149.00, z=-1.380, *P*=0.10). Profiles of contrasting cardiac work but similar AUC METS from two riders are depicted in Figure 2. There was no correlation between total metabolic work (measured as AUC METs) and total cardiac work (measured as AUC HR). A negative correlation was observed between AUC HR and estimated VO_2peak_ (r=-0.51, 95% CI 0.24 to 0.71, *P*=0.001) (Figure 3).

**Figure 2.**
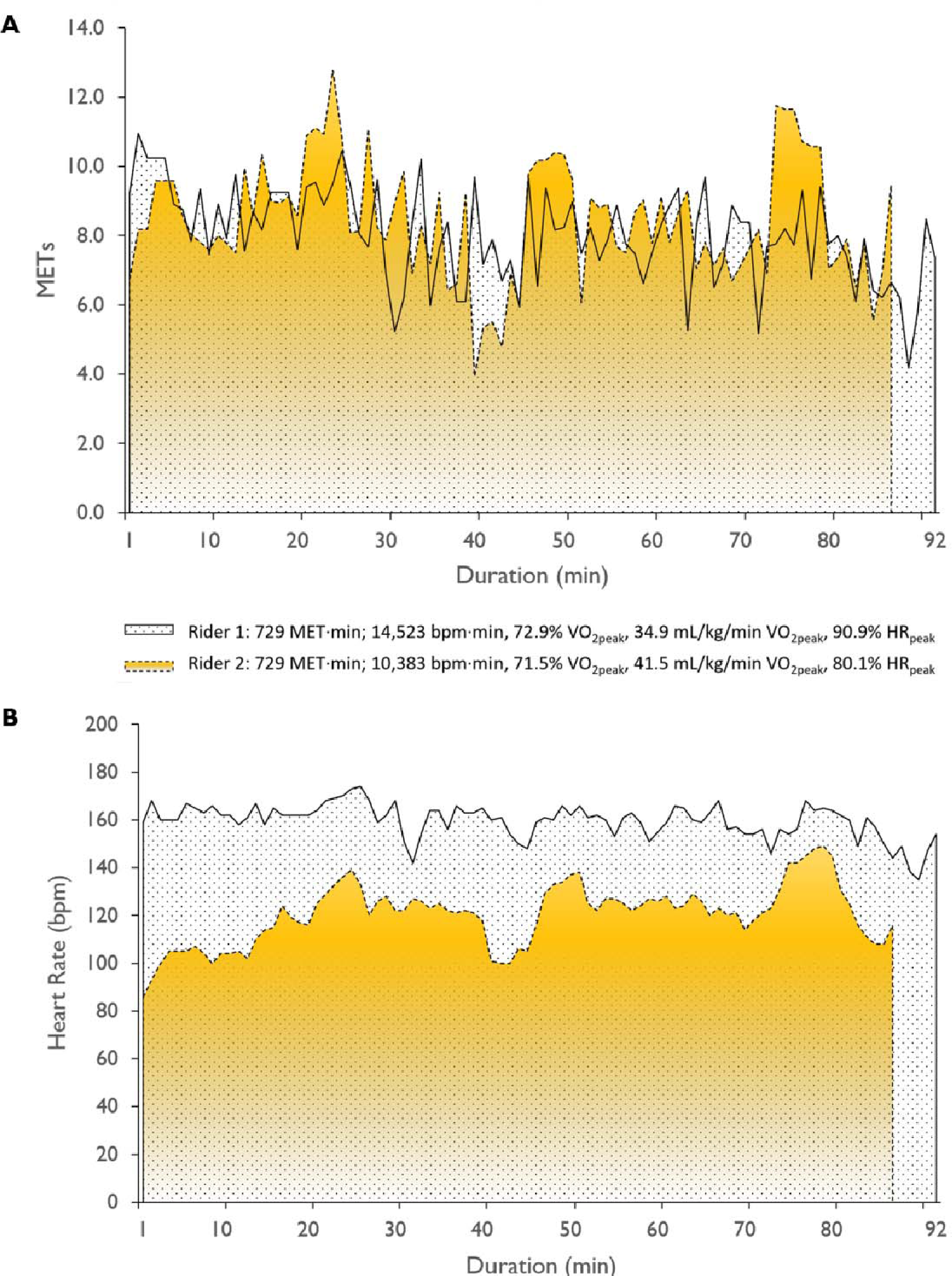
Area under the curve graphs of two male riders (Rider 1, Rider 2; aged between 60-69 years) with equal metabolic burdens (Fig. 2A) but different cardiac burdens (Fig. 2B) for equal distances cycled (42 km).

**Figure 3.**
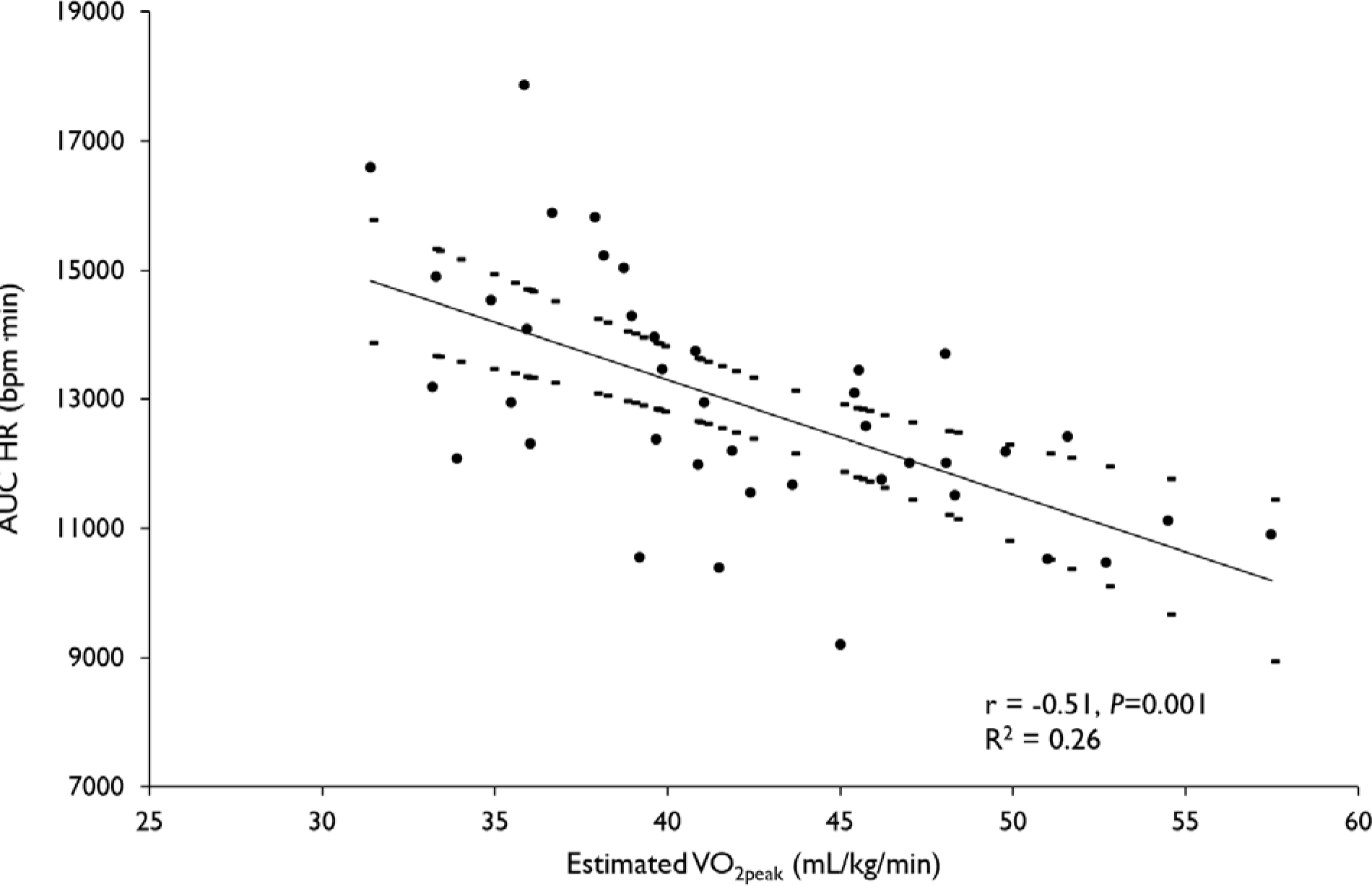
Relationship between area under the curve (AUC) measure of cardiac burden and estimated fitness (VO_2peak_ median = 41.0 mL/kg/min). 95% confidence limits are identified by the dashed line above and below the line of best fit.

To evaluate the impact of fitness on exercise burden, AUC analysis was performed by grouping those above and below the field-based median estimated VO_2peak_ (41.0 mL/kg/min) (Table 3); there was a statistically significant difference between groups for AUC HR and AUC METs even after adjusting for biological sex (*P*=0.001). The more fit subjects produced 16% more energy, but the less fit subjects required 19% more heartbeats during the ride (Table 3).

**Table 3.**
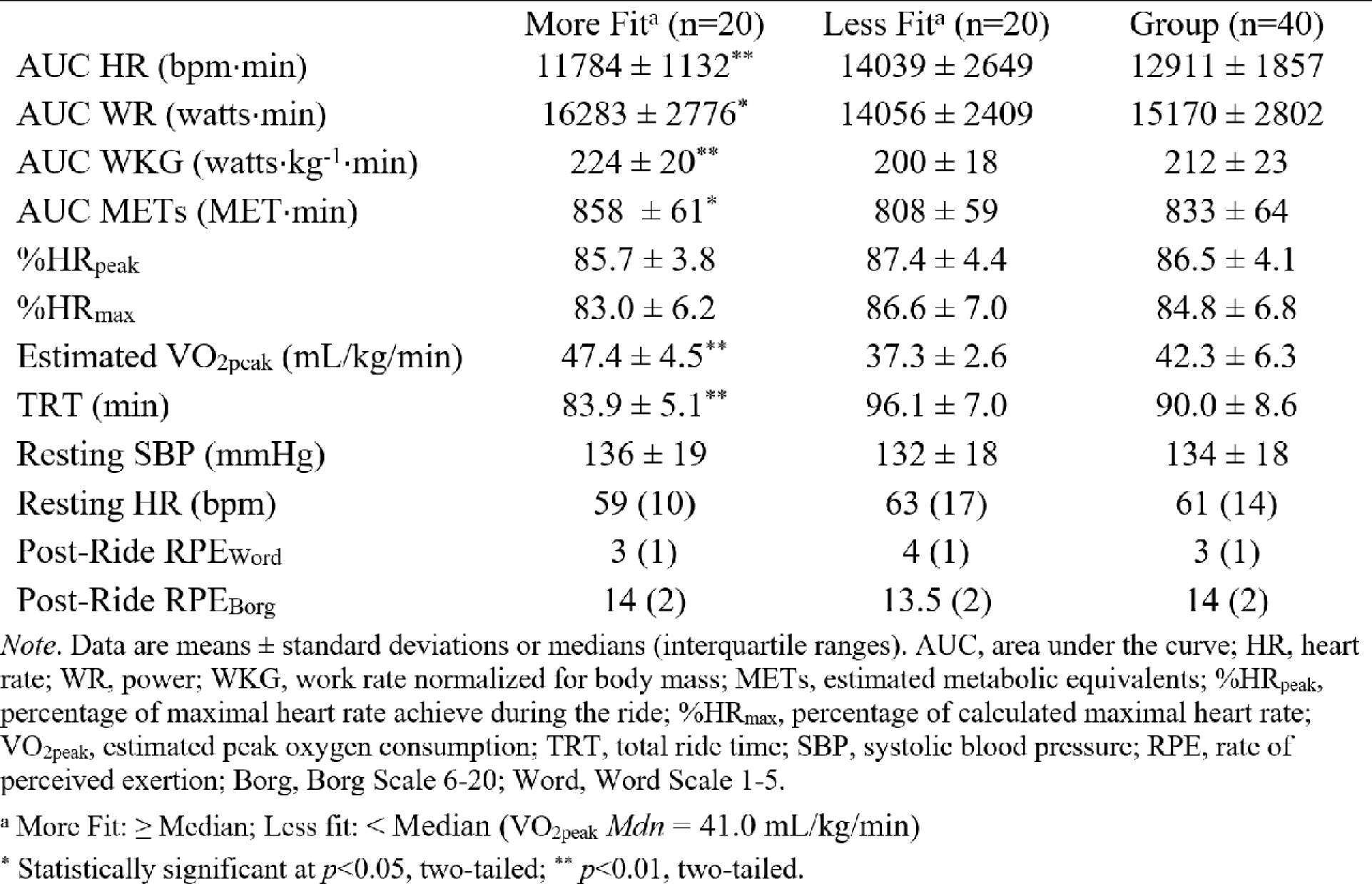
Impact of fitness on key mean and median endpoints of entire ride (42 km).

|  | More Fit<sup>a</sup> (n=20) | Less Fit<sup>a</sup> (n=20) | Group (n=40) |
| --- | --- | --- | --- |
| AUC HR (bpm·min) | 11784 ± 1132** | 14039 ± 2649 | 12911 ± 1857 |
| AUC WR (watts·min) | 16283 ± 2776* | 14056 ± 2409 | 15170 ± 2802 |
| AUC WKG (watts·kg<sup>-1</sup>·min) | 224 ± 20** | 200 ± 18 | 212 ± 23 |
| AUC METs (MET·min) | 858 ± 61* | 808 ± 59 | 833 ± 64 |
| %HR<sub>peak</sub> | 85.7 ± 3.8 | 87.4 ± 4.4 | 86.5 ± 4.1 |
| %HR<sub>max</sub> | 83.0 ± 6.2 | 86.6 ± 7.0 | 84.8 ± 6.8 |
| Estimated VO<sub>2peak</sub> (mL/kg/min) | 47.4 ± 4.5** | 37.3 ± 2.6 | 42.3 ± 6.3 |
| TRT (min) | 83.9 ± 5.1** | 96.1 ± 7.0 | 90.0 ± 8.6 |
| Resting SBP (mmHg) | 136 ± 19 | 132 ± 18 | 134 ± 18 |
| Resting HR (bpm) | 59 (10) | 63 (17) | 61 (14) |
| Post-Ride RPE<sub>Word</sub> | 3 (1) | 4 (1) | 3 (1) |
| Post-Ride RPE<sub>Borg</sub> | 14 (2) | 13.5 (2) | 14 (2) |

*Note.* Data are means ± standard deviations or medians (interquartile ranges). AUC, area under the curve; HR, heart rate; WR, power; WKG, work rate normalized for body mass; METs, estimated metabolic equivalents; %HR<sub>peak</sub>, percentage of maximal heart rate achieved during the ride; %HR<sub>max</sub>, percentage of calculated maximal heart rate; VO<sub>2peak</sub>, estimated peak oxygen consumption; TRT, total ride time; SBP, systolic blood pressure; RPE, rate of perceived exertion; Borg, Borg Scale 6-20; Word, Word Scale 1-5.
<sup>a</sup> More Fit: ≥ Median; Less fit: < Median (VO<sub>2peak</sub> *Mdn* = 41.0 mL/kg/min)
\* Statistically significant at $p < 0.05$ , two-tailed; \*\* $p < 0.01$ , two-tailed.

### Perceived Exertion and Recall Analyses

RPE obtained during exercise (at each landmark; “instantaneous” RPE_Borg_, RPE_Word_) and 30 minutes following the ride (post-ride RPE) varied significantly between participants. Reports of effort were significantly higher if obtained during the ride compared to reports obtained 30 minutes after the ride’s completion, where median RPE_Borg_ during (14.5) was higher than post-ride RPE_Borg_ (14.0, *P*=0.009), similar to that observed for the median RPE_Word_ (3.5) during the ride versus that reported post ride (post-ride RPE_Word_) (3.0; *P*=0.024).

Subjective reports of effort (RPE_Borg_ vs. RPE_Word_) obtained during the ride were well-aligned (r_s_=0.86, 95% CI 0.75 to 0.92, *P*=0.001; Figure 4A) but had less agreement post ride (r_s_=0.54, 95% CI 0.28 to 0.73, *P*=0.001; Figure 4B). In addition, the word descriptor ‘moderate’ exercise intensity was associated with a wide variation of Borg ratings, ranging from 9 to 17 during exercise, and 12 to 16 after exercise. Similarly, ‘vigorous’ exercise was also equated with large ranges of Borg ratings both during the ride [11 to 19] and post ride [13 to 17] (Figure 4). When comparing participant’s word descriptor RPEs during and after exercise, a weak positive correlation was observed (RPE_Word_ vs. post-ride RPE_Word_, r_s_=0.38, 95% CI 0.08 to 0.62, *P*=0.015). However, the RPE_Borg_ and post-ride RPE_Borg_ showed no significant correlation (r_s_=0.31, 95% CI -0.002 to 0.57, *P*=0.051).

**Figure 4.**
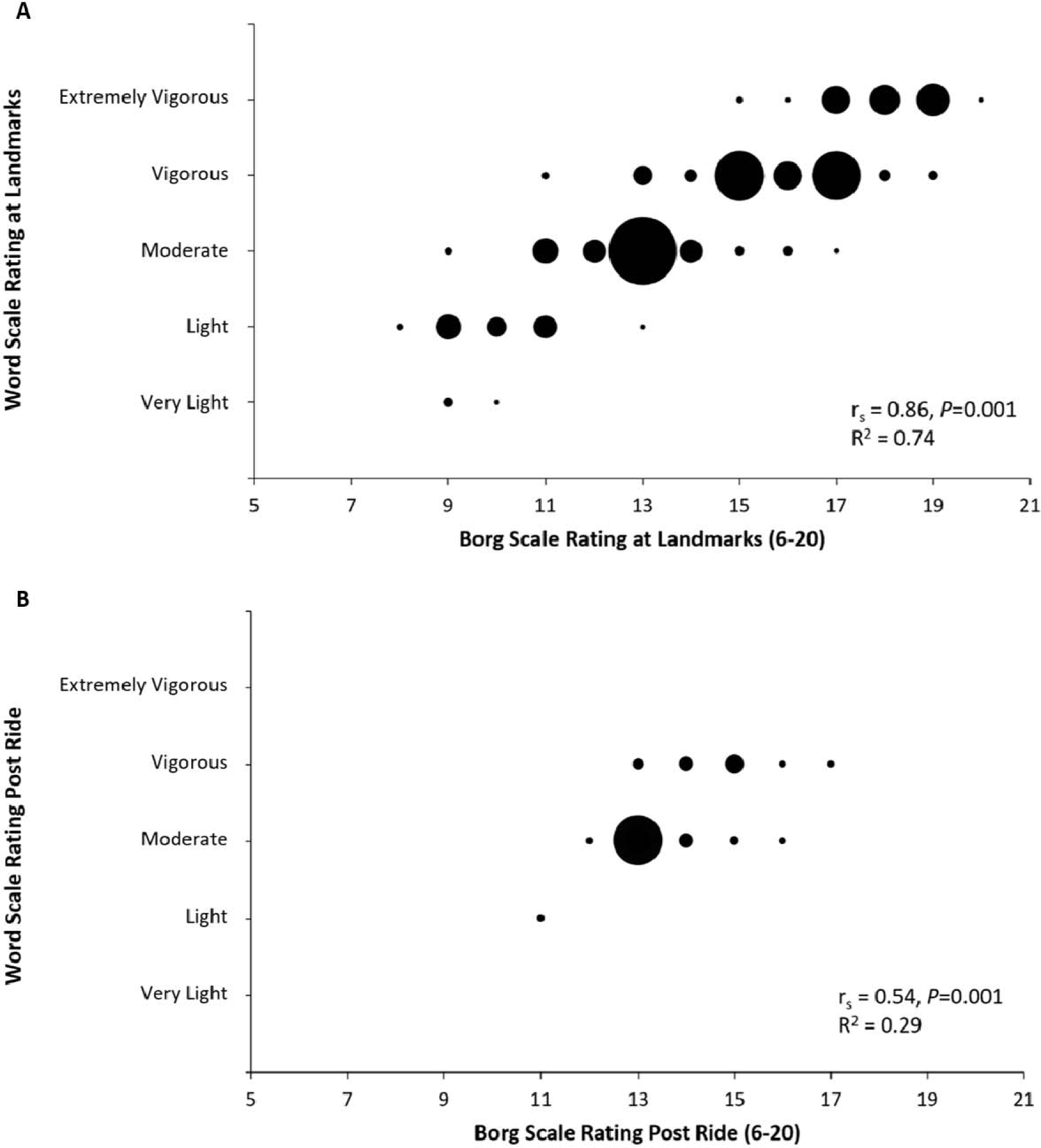
Relationship between Word and Borg 6-20 scale ratings of perceived exertion at landmarks (Fig. 4A) and 30 minutes post ride (Fig. 4B). Dot size represents the frequency of participant responses. Word Scale Ratings: 1=very light, 2=light, 3=moderate, 4=vigorous, 5=extremely vigorous; Borg Scale Ratings: 6=no exertion at all, 9=very light, 11=light, 13=somewhat hard, 15=hard, 17=very hard, 19=extremely hard, 20=maximal exertion.

There were no biological sex differences for post-ride RPE using either perceived exertion scale. The relationship between subjective reports of effort and physiologic data was highly variable, with no association observed between post-ride RPE_Word_ and total AUC HR and AUC MET.

No differences were found between the means of heart rate (Figure 5A, Figure 5C, Figure 5D) and estimated METs (Figure 5B) when riders’ post-ride subjective ratings were grouped based on those who reported ‘moderate’ versus ‘vigorous. However, post-ride RPE _Word_ was negatively correlated with estimated fitness measured as VO_2peak_ (r_s_=-0.32, 95% CI -0.57 to -0.01, *P*=0.04) where higher fitness individuals reported lower post-ride RPE_Word_. Fitness was not significantly correlated with post-ride measures of RPE_Borg_, or “instantaneous” RPE_Word_, or RPE_Borg_.

**Figure 5.**
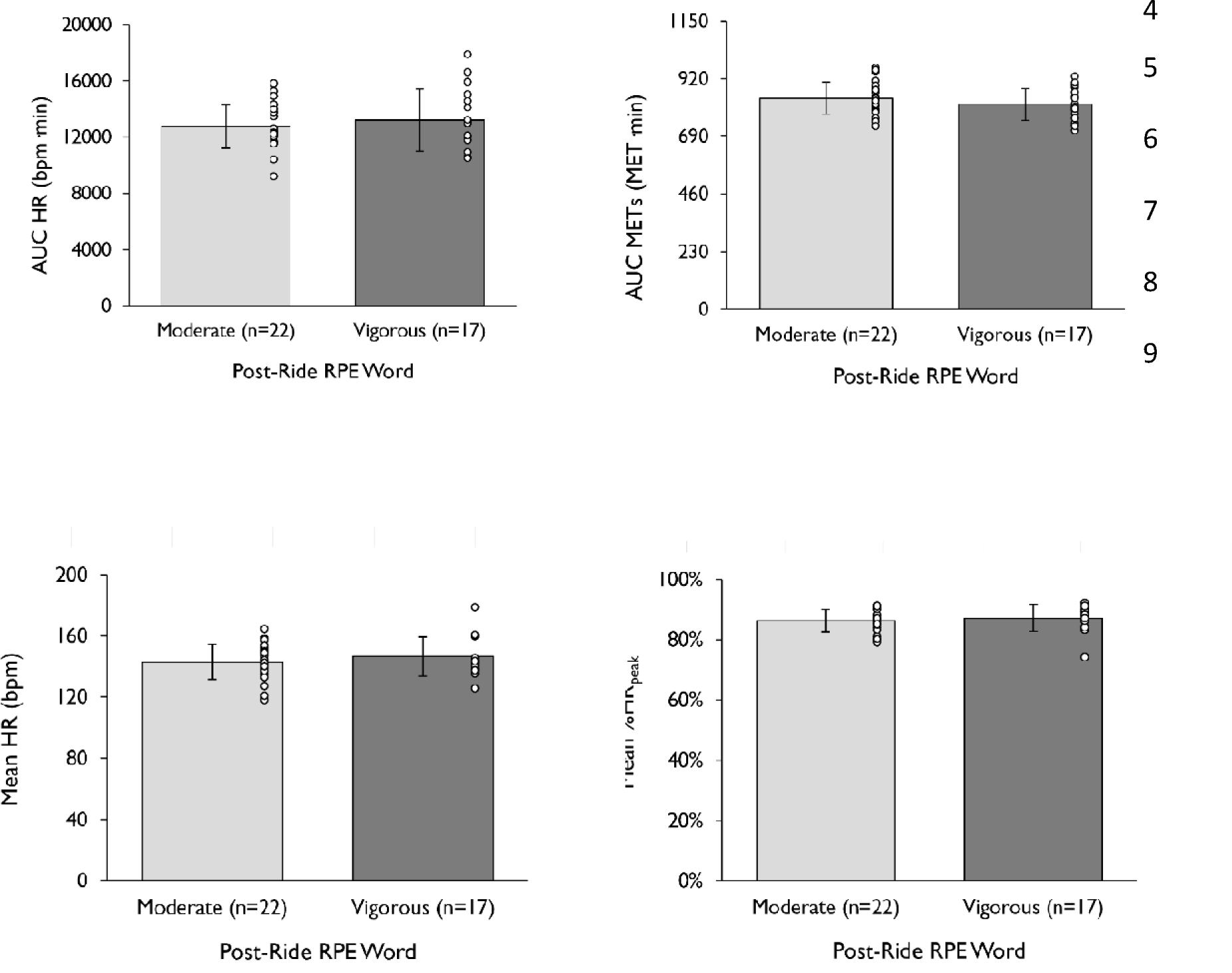
Mean objective measures for post-ride ratings of perceived exertion (RPE) using the Word scale (n=39). Error bars represent standard deviation.

## DISCUSSION

We report novel data describing the relationship between subjective and objective physiological measures of exercise intensity obtained during a typical, field-based, training session in masters endurance athletes. Key findings were: a) subjective reports of effort/intensity using commonly-used verbal descriptors were highly variable between participants and were poorly correlated to objective measures of intensity, especially when reported after exercise; b) measures of exercise ‘burden’ quantified by metabolic energy expenditure, even when performed at the same relative intensity, can be accompanied by significantly different cardiac-specific measures of intensity and are influenced by fitness level. This discordance suggests that a conflation of metabolic and cardiac-specific measures of exercise burden may be inaccurate and misleading.

### Limitations to Self-Reports of Perceived Exertion

While we observed a positive correlation between the two subjective scales of perceived effort at each landmark during exercise, the verbal descriptors of their effort (e.g., “moderate” or “vigorous” effort), were associated with a broad numerical range of the Borg scale (Figure 4), often exceeding the common verbal descriptors described by the ACSM (17). These findings imply that subjective ratings of effort as “moderate” or “vigorous” are not only poorly correlated with actual effort expended but also fail to assess accurately the actual intensity of effort. Our data also demonstrate that individuals with higher fitness levels tend to under-report subjective efforts relative to their true physiological effort, particularly when using traditional categorical classifications. Similar disparate findings were observed for self-reports of effort obtained 30 minutes after ride completion: moderate levels of agreement existed between scales, but there was no agreement between objective measures and subjective descriptors. This was likely due to the wide range of both metabolic and cardiac demands between participants at both ‘moderate’ and ‘vigorous’ intensities. In other words, our data indicate that the reliance on commonly used terms describing effort (e.g., ‘light’, ‘moderate’ or ‘vigorous’) to ascertain exercise intensity, in well-trained athletes, may lead to a substantial error, especially when calculating metabolic exercise intensity (i.e., MET/mins) from compendium data when direct measures of power or velocity are not available. In addition, the mean subjective rating of effort obtained after the ride was associated with a wide range of objective measures; for example, a ‘rating of moderate’ exercise was associated with heart rates ranging from 118 bpm to 165 bpm, with similar disparity observed for METs (7.4 METs to 12.2 METs) and power outputs (112 W to 264 W). Post-exercise subjective recalls of overall effort tended to systematically underestimate the ‘real-time’ effort (i.e., during the ride), and greater intra-individual variability was observed as exercise intensity increased. A high intra-individual variance in ratings of perceived exertion relative to oxygen cost during exercise (12%) has been reported previously (20); collectively, these findings underscore the potential confounding impact of phenotypic variability, including fitness level, on subjective reports of exercise intensity (21, 22).

### Isolating Components of Exercise Burden

A key finding of our study was a discordance of cardiac-specific and global metabolic metrics of exercise intensity amongst athletes of differing fitness levels. As expected, ‘fitter’ athletes had faster ride-completion times but did so performing exercise at a similar relative intensity (%VO_2Peak_ and %HR_Peak_), while at a significantly lower AUC HR (total heartbeats/ride) and higher absolute AUC METs. The higher AUC MET achieved over the same distance covered may reflect greater power outputs achieved during horizontal sections of the ride (vs. gliding), and other phenotypic factors contributing to variability of power output and mechanical efficiency during cycling (23).

Taken together, our data suggest that cardiac burden and perception of effort during vigorous exercise are inversely related to fitness level. These findings may explain prior reports of acute, reversible cardiac dysfunction occurring during prolonged exercise to a greater extent in less experienced participants with lower levels of fitness (24). Observations of increased cardiac risk associated with high intensity exercise may therefore be biased from cohorts of relatively lower fitness levels, who demonstrate a relatively higher cardiac burden of exercise, compared to athletes with superior fitness.

Physical activity recommendations (1, 2) include 150 minutes of moderate to vigorous exercise per week, equivalent to 500 to 1000 MET·min of exercise per week (25). As expected, our participants far exceeded recommended exercise time and MET·min per week by 3 and 6 times, matching previous studies of similar athletes (26, 27). While an estimate of MET·min may infer exercise intensity, in isolation, it can be misleading. Accurate quantification of duration and intensity is required, preferably the integral of each, but reports that include objective measures of intensity remain elusive given technical requirements. We used direct, in-field measures of power and duration to estimate metabolic burden and observed that these measures fail to correlate with self-reports of subjective effort. Even in studies that report MET·min, it is rarely disclosed how such values were obtained but are presumably estimated from self-reports (28–35). MET values may also be estimated from a compendium of physical activity (36, 37), some of which are based on data derived over 60 years ago (38). In one report, exercise intensity was calculated based on data from a questionnaire validated in a clinical, non-athlete population (39).

A strength of our study was its ecological validity, ensuring a self-paced training session which was performed on a common route and training distance, matching the participant’s a priori description of their typical training intensity (range between 69-83%HR_max_ or 56-75%FTP) (40). This approach avoided conditions that would mimic a time trial or race condition that typically elicits exercise intensities beyond 90% VO_2max_ (41). Our data demonstrated that most athletes exercised largely within the ‘vigorous’ zone (17) based on objective measures of heart rate, power output, and METs (17). This is not surprising given the classification of vigorous exercise intensity (>6 METs) was developed for the general population (17) and is far exceeded by well-trained athletes.

### Implications

Acute cardiac events during exercise are associated with vigorous levels of intensity (42), and high exercise heart rates alone have been linked to acute cardiac dysfunction after prolonged exercise (43–46), exercise-related myocardial fibrosis (47–50) and sudden cardiac death (51). However, the paradoxical findings of adverse cardiovascular outcomes linked to long-term cumulative vigorous exercise training remain unresolved. We suggest that efforts to distinguish between the cardiac and metabolic components of exercise will help to provide mechanistic insights into these findings. Ideally, specific metrics of exercise burden should be obtained using direct, objective measures including metabolic and cardiac endpoints. While cardiac minute work (the product of cardiac output and mean arterial pressure) would be a more precise metric than heart rate alone, assessing its constituents would be impractical outside of a laboratory setting. Heart rate can be reliably measured during exercise using wearable technology, providing absolute or relative measures of exercise intensity. While not a complete indicator of cardiac work, it is the most significant factor determining myocardial oxygen cost (52), increasing 3-4 fold during vigorous exercise, whereas systolic blood pressure, a key determinant of left ventricular afterload, may only increase by 1.5-to 2.0-fold.

A more robust measure of cardiac burden would be the integral of heart rate and duration, similar to Banister’s TRIMP method (53) to monitor training intensity (54), especially for determining if there is a threshold effect where cumulative exercise increases the risk for adverse cardiac outcomes. Notwithstanding, simplistic metrics of exercise burden, such as an excess of 1500 cumulative hours of vigorous sport practice, are commonly reported risk factors for atrial fibrillation (AF) (52), yet adults adhering to widely accepted exercise guidelines would surpass numerous thresholds within 12-15 years and are reported to have a *lowered* risk for AF (29).

Moreover, there is wide discrepancy in the cumulative hours associated with AF risk [1500 hours to 4500 hours of exercise (52–54, 55)] or metabolic burden [1900 MET·min per week (35) to 5000 MET·min per week (34)], which may reflect an overly simplistic approach when quantifying total exercise burden, especially when based on self-reports (55). Our data suggest that subjective self-reports obtained soon after exercise fail to accurately reflect objective endpoints obtained during exercise, are less precise than a rating of perceived exertion (Borg) and show diminishing accuracy over time. Therefore, the reliance on subjective self-reports to estimate exercise ‘burden’ simply based on ’hours of exercise’ or its metabolic cost (MET·min), may be inaccurate and misleading indicators of exercise burden.

### Limitations

Our study has limitations. Effort was made to ensure our participants were typical recreational, sub-elite cyclists based on their training history. While recall bias was possible, their training histories were determined by questionnaire and verified by digital records of training history through mobile applications. All field data were subject to varying environmental conditions (e.g., wind, temperature) that may have influenced physiological and perceptual efforts. We were limited to estimations of METs based on power outputs given institutional COVID-19 restrictions that precluded direct gas-exchange measures in the field or laboratory; estimates of VO_2peak,_ derived from relationships between peak power and in-field peak heart rate, may have led to error. Quantifying metabolic and cardiac burden plus subjective effort during a non-weight-bearing activity such as cycling may introduce error because external work at times can be zero or nominal given the ability to glide, reflected by the modest correlation between mean heart rate and mean power (r_s_=0.34). Consequently, our findings may not be generalizable to other weight-bearing endurance activities such as running, where exercise intensity is less variable. Lastly, we recognize the limitations of using AUC HR as a measure of “cardiac burden”, but additional hemodynamic measures (cardiac output, ambulatory systolic blood pressure) were not feasible.

## Conclusion

This study demonstrated a discordance between the overall metabolic and cardiac burden of exercise. Less fit endurance athletes completed a fixed training distance with a lower metabolic burden but at a higher cardiac burden, despite performing exercise at a similar relative intensity. Post-exercise subjective reports of exercise intensity were variable at high levels of exercise intensity and did not align with objective, physiological measures of effort. These findings demonstrate the complexities of assessing exercise burden and suggest that caution is warranted when interpreting studies that report simplistic, subjective reports of effort used to estimate exercise intensity, and in particular, relate exercise history to cardiovascular outcomes. The use of physiological endpoints is advised when designing studies that consider the impact of long-term exercise training and its relationship to the risks of adverse cardiac outcomes.

## FUNDING

Funding for this study was provided by the Canadian Institutes for Health Research (CIHR; Operating Grant 130477) and from the Heart & Stroke/Richard Lewar Centres of Excellence.

## Data Availability

All data produced in the present study are available upon reasonable request to the authors.

## STRUCTURED GRAPHICAL ABSTRACT

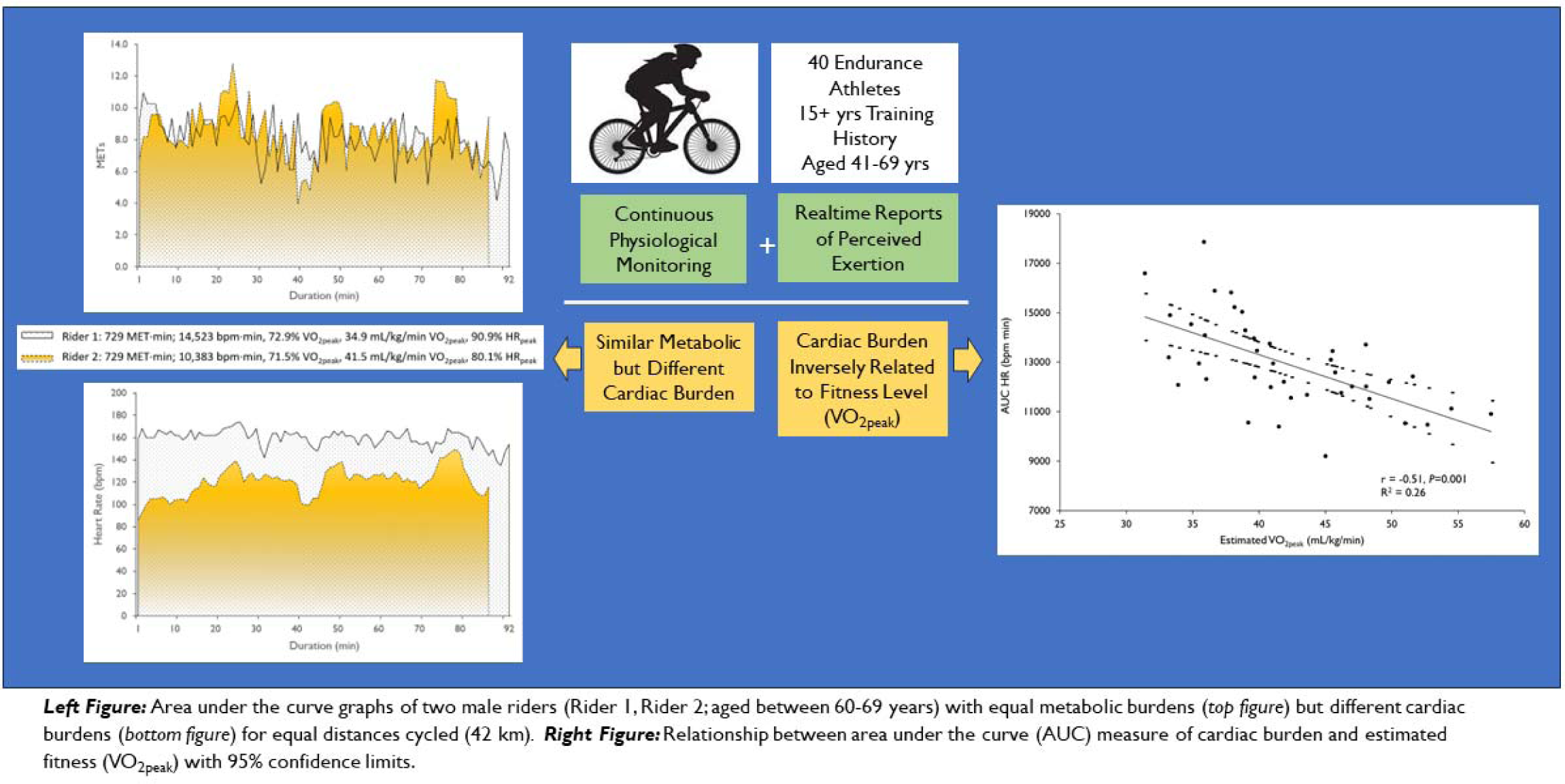

## STRUCTURED GRAPHICAL ABSTRACT TEXT

### Key Questions

Are subjective ratings of exercise intensity from endurance athletes congruent with objective measures obtained in the field, and is it important to distinguish between the cardiac and metabolic burden of exercise?

### Key Findings

Post-exercise subjective reports of exercise intensity were variable at high levels of exercise intensity and did not align with objective, physiological measures of exertion. In addition, there is a discordance between metabolic and cardiac burden during intensive exercise, with cardiac burden being inversely related to fitness level.

### Take Home Message

Estimates of exercise intensity from self-reports are highly variable and a conflation of metabolic and cardiac-specific measures of exercise burden may be misleading when considering the impact of long-term exercise training and its relationship to the risks of long-term adverse cardiac outcomes.

**Suppl Table 1.** Key endpoints at each landmark (n=40).

| LM Gradient (%) | LM1-M<br>0.9 | LM2-H<br>6.7 | LM3-L<sup>a</sup><br>0.2 | LM4-H<br>1.5 | LM5-M<br>1.2 | LM6-VH<br>-0.4 |
| --- | --- | --- | --- | --- | --- | --- |
| Absolute Objective Measures |  |  |  |  |  |  |
| HR (bpm) | 143 ± 13* | 160 ± 12 | 131 ± 17* | 157 ± 11 | 145 ± 13* | 159 ± 12 |
| WR (watts) | 185 ± 48* | 236 ± 65* | 131 ± 46* | 204 ± 55 | 158 ± 50* | 206 ± 59 |
| WR/body mass (W/kg) | 2.6 ± 0.5* | 3.3 ± 0.7* | 1.8 ± 0.6* | 2.8 ± 0.5 | 2.2 ± 0.6* | 2.9 ± 0.6 |
| METs | 10.0 ± 1.5* | 12.1 ± 2.0* | 7.7 ± 1.7* | 10.8 ± 1.7 | 8.8 ± 1.7* | 10.8 ± 1.9 |
| Relative Objective Measures |  |  |  |  |  |  |
| %HR<sub>max</sub> | 84 ± 8* | 94 ± 7 | 77 ± 10* | 92 ± 6 | 85 ± 7* | 93 ± 7 |
| %HR<sub>peak</sub> | 86 (7)* | 98 (3) | 80 (11)* | 95 (4) | 88 (7)* | 96 (5) |
| %MET<sub>peak</sub> | 84 ± 11* | 101 ± 6* | 64 ± 4* | 90 ± 9 | 73 ± 8* | 90 ± 7 |
| Relative Subjective Measures |  |  |  |  |  |  |
| RPE<sub>Borg</sub> (6-20) | 13 (2)* | 17 (1)* | 10 (2)* | 16.5 (2)* | 13 (2)* | 18 (2) |
| RPE<sub>Word</sub> (1-5) | 3 (2)* | 4 (1) | 2 (0)* | 4 (1)* | 3 (0)* | 5 (1) |

*Note.* Data are means ± standard deviations or medians (interquartile ranges). Prescribed efforts: M, medium; H, high; L, low; VH, very high. LM, landmark; HR, heart rate; WR, power; W, watts; METs, estimated metabolic equivalents; %HR<sub>max</sub>, percentage of calculated maximal heart rate; %HR<sub>peak</sub>, percentage of peak heart rate; %MET<sub>peak</sub>, percentage of estimated peak metabolic equivalents; RPE, rating of perceived exertion; Borg, Borg Scale; Word, Word Scale.
<sup>a</sup> All variables at LM3 were statistically significant from all other LMs at $p < 0.05$ . For simplicity, significant symbols were not shown.
\* Statistically significant landmark from LM6 at $p < 0.05$ .

## Notes

### Competing Interest Statement

The authors have declared no competing interest.

### Funding Statement

Funding for this study was provided by the Canadian Institute for Health Research (CIHR), Operating Grant 130477 and from the Heart & Stroke/Richard Lewar Centres of Excellence.

### Author Declarations

Approval for this study was provided by the University of Toronto Health Sciences Research Ethics Board (RIS RIS Human Protocol 39300).

